# Neonatal Anti-seizure Medication Exposure and Long-term Neurological Outcomes

**DOI:** 10.64898/2026.02.02.26345337

**Authors:** Markus Leskinen, Volmar Kaipainen, Mine Öğretir, Marjo Metsäranta, Matti Hero, Samuli Rautava, Miika Koskinen

**Author notes:** Address correspondence to: Markus Leskinen, Department of Neonatology, New Children’s Hospital, P.O. Box 347, 00029 HUS, Finland.

## Abstract

**Objective:** There is limited evidence for Anti-seizure medication (ASM) safety and long-term outcomes in neonates. Phenobarbital is the only ASM accepted by FDA for neonatal use, but it has been linked with adverse effects. Other ASMs are used off-label in neonates. We wanted to examine links between neonatal ASM exposure and later neurological outcomes.

**Methods:** We performed a retrospective observational study of 18,548 infants in intensive care, examining links between ASM exposure and neurological outcomes over follow-up period of median 4.5 years. The clinical data included comprehensive maternal, perinatal, and medication data. Multivariable cause-specific Cox models were used to estimate hazard ratios (HRs) for phenobarbital, levetiracetam, midazolam, and fosphenytoin exposure.

**Results:** Exposure to the median cumulative dose of phenobarbital was associated with increased HR for epilepsy (HR 1.35; 95% CI, 1.11–1.62) visual impairment (HR 1.20; 95% CI, 0.99–1.45), and intellectual disability (HR 1.18; 95% CI, 0.99–1.41). In contrast, levetiracetam was associated with smaller risk increases for cerebral palsy (HR 1.13; 95% CI, 1.03–1.23), epilepsy (HR 1.14; 95% CI 1.05–1.24) and visual impairment (HR 1.18; 95% CI 1.11–1.26).

We did not find evidence of a dose-dependent phenobarbital effect, but increased maximum concentrations were associated with elevated hazard ratios for cerebral palsy (HR 1.48; 95% CI, 1.07–2.06) and epilepsy (HR 1.64; 95% CI, 1.14–2.35).

**Conclusion:** The results align with previous findings linking phenobarbital to neurodevelopmental harm and emphasize the need for its cautious use in neonates. Levetiracetam had more favorable safety profile.

## Introduction

Neonatal intensive care relies heavily on pharmacological interventions and polypharmacy is common. Due to low patient numbers and inherent challenges of conducting pharmacological research in infants, many of the used medications are either off-label^1,2^ or lack robust evidence regarding their long term safety and efficacy. Recently, the utilization of clinical real-world data in pharmaceutical research has garnered increasing interest for its use in evaluating the effectiveness and safety of medication, particularly in situations where traditional large-scale clinical trials are difficult to design or are otherwise challenging.

Seizures are common emergency in Neonatal intensive care units (NICUs) affecting 1–5 infants per 1000 live births. In high income countries the most common etiologies include hypoxic-ischemic encephalopathy, ischemic stroke, and intracranial hemorrhage. Prompt treatment with anti-seizure medication (ASM) is critical to minimize brain injury, but evidence regarding the optimal choice of ASM and their long-term effects remains limited.^3,4^

Currently phenobarbital is the only ASM approved by the FDA for use in neonates and remains the first-line treatment in most settings.^4,5^ Other anti-seizure medications, such as fosphenytoin, levetiracetam, and midazolam are used off-label in this population. There are limited number of randomized controlled trials favoring phenobarbital over phenytoin and levetiracetam^6,7^ whilst some studies suggest comparable efficacy between phenobarbital and levetiracetam.^8,9^ Despite its widespread use, phenobarbital has been linked with adverse effects in infants, with hypotension and respiratory depression among the most frequently reported acute complications.^3^ Evidence regarding its long-term effects is sparse typically limiting to the first two years of life. Experimental animal studies have reported concerning findings, including neuronal apoptosis, altered synaptic maturation, and changes in long term behavior, learning and memory,^10–12^ but currently there are limited data from humans.

Postnatal exposure to phenobarbital has been associated with cognitive decline in humans^13,14^ but due to coexisting hypoxic-ischemic injury and/or the effects of repeated seizures on many of the infants receiving phenobarbital, causality is difficult to assess. Phenobarbital was also previously used to treat febrile seizures, and in this setting it depressed cognitive performance of children randomized to receive phenobarbital.^15^

Levetiracetam, by contrast, demonstrates a favorable safety margin, with most reported adverse effects considered clinically manageable.^16,17^ One retrospective study found worse neurodevelopmental outcomes in infants treated with phenobarbital than levetiracetam.^18^ The favorable safety profile has contributed to the increasing off-label use of levetiracetam for neonatal seizures, but its optimal use has remained elusive.^19^

Fosphenytoin, historically a second line therapy for treating neonatal seizures, has seen a decline in use over recent years.^20^ The adverse effects of fosphenytoin include arrhythmia, hypotension and central neural system depression.^6^ In animal studies it has also caused apoptosis and synaptic disruption.^11,12^

Midazolam is typically reserved as a third-line therapy for refractory seizures, yet most studies assessing its efficacy in neonates are limited by small sample sizes and inconsistent findings.^21,22^ As a potent benzodiazepine, midazolam carries risks of sedation and need for mechanical ventilation,^23^ and increased risk for poor early neurological outcome in patients sedated with midazolam as compared with morphine has been reported.^24^

Against this backdrop, we aimed to investigate the long-term neurological outcomes associated with neonatal ASM exposure. By linking comprehensive real-world NICU data on ASM administration with registry-based follow-up information concerning major neurologic morbidities—including cerebral palsy, epilepsy, autism spectrum disorders, and developmental problems—we were able to assess population-level clinical outcomes extending well beyond the common two-year follow-up period.

## Materials and Methods

### Study design and population

This retrospective observational study included all the infants admitted to the neonatal intensive care unit of Helsinki University Hospital (HUS), a level III/IV NICU with a catchment area of approximately 18,000 deliveries per year, between January 1, 2009, and March 31, 2024. Following the predefined exclusion criteria, the final follow-up cohort consisted of 18,548 infants. All analyses were conducted using a secure research platform (HUS Acamedic), which complies with national and EU General Data Protection Regulation. HUS Helsinki University Hospital approved the study protocol (HUS/617/2025). Individual consent was not required for this register study.

### Clinical data

The dataset included maternal and neonatal demographic data, birth details, administered medications, and diagnoses both from the initial hospitalization and long-term inpatient and outpatient encounters within HUS Helsinki University Hospital.

Medications were identified with ATC-codes or generic names. Weight-adjusted cumulative doses were calculated from medication administration data. Medication concentrations were retrieved from laboratory data. Early diagnoses were identified by ICD-10 codes entered for the NICU stay. Birth weight, gestational age, patient sex and mode of delivery were extracted from birth records.

Hospital discharge was defined as the end date of the initial hospitalization record, after which cumulative medication doses were computed. The follow-up times were calculated from discharge to the diagnosis date, death or to the date of last contact with the patient (censoring time).

A total of 34 patients with missing data in key variables such as birth weight, mode of delivery, gender, or maternal identity were excluded from the final dataset.

Data were queried from the HUS Datalake which aggregates data from multiple patient information systems. In the years 2009-2020 the NICU data were recorded in Centricity Critical Care Clinisoft (GE Healthcare, Chicago, IL, USA), prenatal care and delivery information in Obstetrix (Siemens, Sweden) and pediatric care in Uranus (CGI, Finland). In 2020, HUS transitioned to the EPIC patient information system (EPIC Systems, Verona, WI, USA)

### Outcomes

Neurological outcomes were determined using ICD-10 codes from follow-up visits. The outcomes assessed included cerebral palsy, epilepsy, intellectual disability, autism spectrum disorders and sensorineural defects consisting of visual impairment and hearing impairment. The ICD-10 codes and the number of patients for each outcome are shown in Table 1. Due to the small number of patients the association between ASM exposure and autism spectrum disorder or hearing impairment could not be evaluated (Table 2).

**Table 1:**
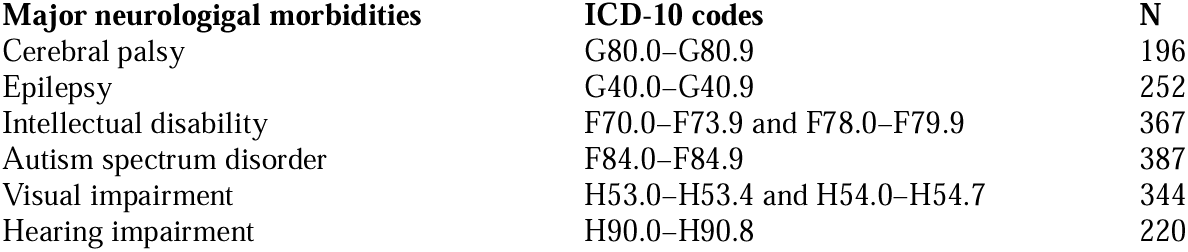
Table listing neurological outcomes studied in the paper, their ICD-10 codes and number of cases. Most common outcomes were autism spectrum disorder with 387 cases and intellectual disability with 367 cases.

| Major neurologigal morbidities | ICD-10 codes | N |
| --- | --- | --- |
| Cerebral palsy | G80.0–G80.9 | 196 |
| Epilepsy | G40.0–G40.9 | 252 |
| Intellectual disability | F70.0–F73.9 and F78.0–F79.9 | 367 |
| Autism spectrum disorder | F84.0–F84.9 | 387 |
| Visual impairment | H53.0–H53.4 and H54.0–H54.7 | 344 |
| Hearing impairment | H90.0–H90.8 | 220 |

**Table 2:** Table showing patient demographics, early diagnosis and later neurological morbidities in all study patients and patients exposed to phenobarbital, levetiracetam, midazolam, and fosphenytoin.

|  | All | Phenobarbital | Levetiracetam | Midazolam | Fosphenytoin |
| --- | --- | --- | --- | --- | --- |
| N | 18548 | 336 | 136 | 229 | 37 |
| **Patient demographics** |  |  |  |  |  |
| Male | 10 261 (55.3 %) | 182 (54.2%) | 71 (52%) | 133 (58.1%) | 22 (60 %) |
| Birth weight (Kg, Mean, SD) | 3.00 ± 0.93 | 3.17±0.91 | 3.13±0.79 | 3.12 ± 0.77 | 3.17 ± 0.63) |
| Gestational age (Weeks, Mean, SD) | 37.4 ± 3.65 | 38.4 ± 3.9 | 38.6 ± 3.25 | 38.7 ± 2.95 | 38.9 ± 2.48 |
| Vaginal delivery | 9892 (53.3 %) | 168 (50.0%) | 55 (40 %) | 129 (56.3 %) | 19 (51%) |
| Hospitalization time (Days, Median, IQR) | 5.4 (3.4-12.7) | 14.1 (9.0-28.7) | 16.0 (10.7-27.0) | 27.5 (19.2-53.8) | 13.6 (10.0-20.7) |
| Cumulative dose (mg/kg, Median, IQR) |  | 28.4 (20.0-37.3) | 98.6 (30.4-203.6) | 2.5 (0.7-6.9) | 23.8 (20.6-31.3) |
| **Early diagnoses (ICD-10)** |  |  |  |  |  |
| Severe birth asphyxia (P21.0) | 362 (2.0%) | 124 (36.9%) | 52 (38 %) | 34 (14.8) | 9 (24%) |
| Mild and moderate birth asphyxia (P21.1) | 984 (5.3%) | 40 (11.9%) | 12 (9 %) | 5 (2.2) | 4 (11%) |
| Cerebral ischemia (P91.08) | 80 (0.4%) | 59 (17.6%) | 42 (31%) | 9 (3.9) | 17 (46%) |
| Mild hypoxic-ischemic encephalopathy (P91.00) | 82 (0.4%) | 14 (4.2%) | < 3 | < 3 | < 3 |
| Moderate hypoxic-ischemic encephalopathy (P91.01) | 95 (0.5%) | 54 (16.1%) | 16 (12%) | 11 (4.8) | 4 (11%) |
| Severe hypoxic-ischemic encephalopathy (P91.02) | 47 (0.3%) | 44 (13.1%) | 32 (24%) | 19 (8.3) | 7 (19%) |
| Intraventricular hemorrhage, grade 1 (P52.0) | 223 (1.2%) | 5 (1.5%) | < 3 | 3 (1.3) | < 3 |
| Intraventricular hemorrhage, grade 2 (P52.1) | 128 (0.7%) | 3 (0.9%) | < 3 | 5 (2.2) | < 3 |
| Intraventricular hemorrhage, grade 3 and grade 4 (P52.2) | 108 (0.6%) | 19 (5.7%) | 10 (7%) | 4 (1.7) | < 3 |
| **Later morbidities (ICD-10)** |  |  |  |  |  |
| Cerebral palsy (G80-G80.9) | 196 (1.1%) | 85 (25.3%) | 53 (38%) | 26 (11.4) | 14 (38%) |
| Epilepsy (G40-G40.9) | 252 (1.4%) | 57 (17.0%) | 36 (27%) | 19 (8.3) | 10 (27%) |
| Intellectual disability (F70.0-F73.9 and F78.0-F79.9) | 367 (2.0%) | 49 (14.6%) | 25 (18%) | 28 (12.2) | 6 (16%) |
| Autism spectrum disorder (F84-F84.9) | 387 (2.1%) | 13 (3.9%) | 6 (4%) | 9 (3.9) | < 3 |
| Visual impairment (H53-H54 and H54.0-H54.7) | 344 (1.9%) | 42 (12.5%) | 24 (18%) | 18 (7.9) | 7 (19%) |
| Hearing impairment (H90-H90.8) | 220 (1.2%) | 15 (4.5%) | 8 (6%) | 15 (6.6) | < 3 |

Data are n (%) or mean (SD). If there were less than 3 patients, <3 is shown instead of exact number.

### Statistical Analysis

Multivariable cause-specific hazard Cox models were employed to examine the associations between exposure to phenobarbital, levetiracetam, midazolam and fosphenytoin and the risk of cerebral palsy, epilepsy, visual impairment and intellectual disability. The time-to-event was defined using the follow-up time calculated earlier for each patient. Four separate models were constructed for each medication-outcome pair. Potential dose–response relationships were examined by including the medication use—defined as the total cumulative dose during hospitalization—as a continuous variable in the Cox models. Sensitivity analyses were conducted to account for possible nonlinear associations between medication exposure and outcome hazard using restricted cubic splines.

The covariates adjusted for in the models for each outcome were determined by prior knowledge of relevance. These included gestational age, birth weight, mode of birth, gender, intraventricular hemorrhage (IVH), neonatal hypoxic-ischemic encephalopathy (HIE), birth asphyxia, stroke, hospitalization time (age at discharge) and retinopathy of prematurity. IVH and HIE were categorized into severe versus moderate/mild forms, and mode of delivery was categorized as vaginal birth or cesarean section.

### Analysis of multiple exposures

To account for 30% of patients receiving more than one ASM during their hospitalization, each model was adjusted for co-exposure to other ASMs. Additionally, since phenobarbital is used as the first-line ASM, exposure to multiple treatments may indicate poor response and more difficult clinical condition. This in turn may lead to residual confounding when comparing phenobarbital to other ASMs. Thus, to explore whether multiple ASM use was associated with poorer outcomes, confounder-adjusted cumulative incidence functions were estimated between patients exposed to phenobarbital and patients exposed to phenobarbital and other ASMs using direct standardization.^25^

### Phenobarbital blood concentration measurements

For patients with phenobarbital serum concentration measurements, we explored whether the maximum concentration level was associated with adverse outcomes by fitting additional models for each outcome. The models were adjusted to IVH, HIE and stroke only due to the smaller sample size and number of outcomes in the patient subgroup.

### Sensitivity Analysis

To evaluate the robustness of our modelling assumptions and data selection procedures, two sensitivity analyses were conducted. Firstly, follow-up was restricted to 95th percentile of event times for each outcome to examine whether late diagnoses potentially biased our results. Secondly, cumulative doses were calculated only up to 28 days from birth, with follow-up commencing from that time point. This was done to assess whether large cumulative dose outliers accumulating for patients with longer hospitalization periods influenced our results.

To account for within-family correlation due to siblings in the data, cluster-robust standard errors were employed. The proportional hazards assumption was assessed visually with Schoenfeld residual plots.^26^ Variables that violated the assumption were either stratified with or their coefficients were allowed to vary with time using a step function.

All statistical analyses were performed using R version 4.5.0 (R Statistical Computing Foundation, Vienna, Austria) and packages *survival, rms, cmprsk* and *adjustedCurves.* All statistical tests were two-sided and a P-value <0.05 was considered significant.

ChatGPT Version 1.2025.350 and Microsoft Copilot were used in order to refine the language of the work.

## Results

### Patients and identification

A total of 24,145 infants admitted to the HUS NICU during the study period were initially included. Infants who died before discharge, had missing data, were residents outside the HUS catchment area, had no follow-up information or were born after 2023 were excluded, leaving a final analytic cohort of 18,548 infants (Figure 1).

**Figure 1:**
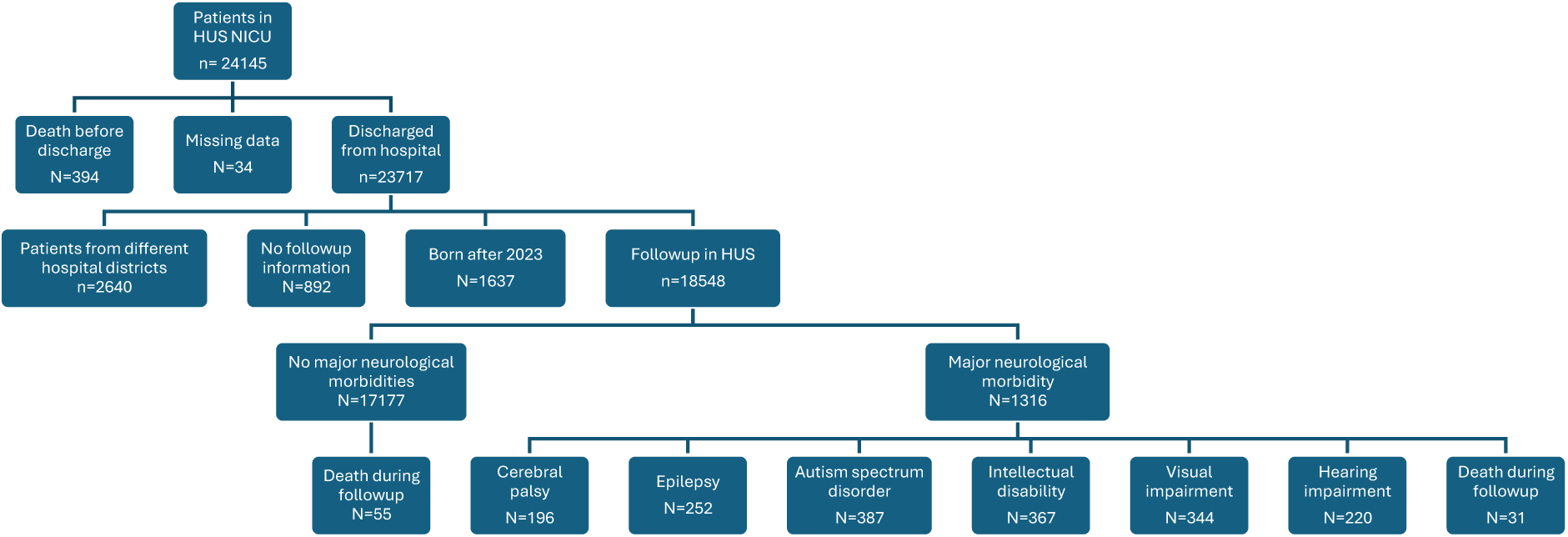
Patient flowchart. Alt Texts: Flowchart showing patient selection, reasons for exclusion, and numbers of patients in each group.

The demographic and clinical characteristics of the cohort are summarized in Table 2. The median follow-up times calculated with reverse Kaplan-Meier estimators were 4.5 years (average IQR: 1.6 – 9.2 years) for each diagnosis, with the follow-up times varying slightly depending on the outcome. During the follow-up period, 196 cases of cerebral palsy, 252 of epilepsy, 367 of intellectual disability, 387 of autism spectrum disorder, 344 of visual impairment, and 220 of hearing impairment were identified. A total of 515 subjects received any ASM with 370 subjects receiving one, 79 subjects two, 54 subjects three and 12 subjects all four ASMs. The use of phenobarbital only (n = 193) was most common, followed by midazolam only (n = 171), both phenobarbital and levetiracetam (n = 64) and a combination of phenobarbital, levetiracetam and midazolam (n = 33).

### Adjusted Hazard Ratios

Exposure to the median cumulative dose of phenobarbital was associated with increased hazard ratio for epilepsy (HR 1.35; 95% CI, 1.11–1.62, p = 0.002) visual impairment (HR 1.20; 95% CI, 0.99–1.45, p = 0.06), and intellectual disability (HR 1.18; 95% CI, 0.99–1.41, p = 0.06). Increased hazard ratio for cerebral palsy was not statistically significant (HR 1.13; 95% CI, 0.83–1.53, p = 0.44) (Table 3).

**Table 3:** Table showing hazard ratios to cerebral palsy, epilepsy, visual impairment and intellectual disability in patients exposed median cumulative dose of phenobarbital, levetiracetam, midazolam, or fosphenytoin. Highest statistically significant hazard ratio was 1.35 for epilepsy in patients exposed to phenobarbital.

|  | Cerebral Palsy<sup>1</sup> | Epilepsy<sup>1</sup> | Visual Impairment<sup>2</sup> | Intellectual Disability<sup>3</sup> |
| --- | --- | --- | --- | --- |
| **Phenobarbital** | 1.13 [0.83, 1.53]<br>P = 0.44 | 1.35 [1.11, 1.62]<br>P = 0.002 | 1.20 [0.99, 1.45]<br>P = 0.06 | 1.18 [0.99, 1.41]<br>P = 0.06 |
| **Levetiracetam** | 1.13 [1.03, 1.23]<br>P = 0.006 | 1.14 [1.05, 1.24]<br>P = 0.002 | 1.18 [1.11, 1.26]<br>P <0.0001 | 1.08 [0.98, 1.19]<br>P = 0.14 |
| **Midazolam** | 1.03 [0.89, 1.20]<br>P = 0.74 | 1.06 [0.97, 1.16]<br>P = 0.20 | 1.00 [0.85, 1.18]<br>P = 0.97 | 1.09 [1.02, 1.16]<br>P = 0.01 |
| **Fosphenytoin** | 0.900 [0.36, 2.25]<br>P = 0.81 | 1.62 [0.82, 3.21]<br>P = 0.14 | 1.87 [0.95, 3.69]<br>P = 0.07 | 1.10 [0.49, 2.43]<br>P = 0.82 |
| <sup>1</sup> Adjusted for gestational age, birth weight, hospitalisation time, mode of delivery, gender, IVH, HIE, stroke and birth asphyxia.<br><sup>2</sup> Adjusted for gestational age, birth weight, hospitalisation time, mode of delivery, gender, IVH, HIE, stroke, birth asphyxia and retinopathy of prematurity.<br><sup>3</sup> Adjusted for gestational age, birth weight, hospitalisation time, mode of delivery, gender, IVH, HIE and birth asphyxia. |  |  |  |  |

Hazard ratios are reported for patients receiving the median positive cumulative dose for each medication, when the reference is a patient with no exposure. No statistical corrections for multiple testing were applied to the reported P-values.

The adjusted hazard ratios associated with exposure to the median cumulative dose of levetiracetam were on average smaller, but more precise, than those observed for phenobarbital (Table 3). Levetiracetam was associated with increased hazard ratios for cerebral palsy (HR 1.13; 95% CI, 1.03–1.23 *p* = 0.006), epilepsy (HR 1.14; 95% CI, 1.05–1.24; *p* = 0.002) and visual impairment (HR 1.18; 95% CI, 1.11–1.26; *p* < 0.001) The hazard ratio for intellectual disability was not statistically significant (HR 1.08; 95% CI, 0.98–1.19; p = 0.14).

Midazolam administration demonstrated fewer adverse effects than levetiracetam and phenobarbital. Midazolam exposure was significantly associated with intellectual disability (HR 1.09; 95%, CI 1.02–1.16, p = 0.01). The HRs for cerebral palsy was 1.03 (95%, CI 0.89–1.20, p = 0.74), epilepsy 1.06 (95% CI, 0.97–1.16, p = 0.20), and visual impairment 1.00 (95% CI, 0.85–1.18. p = 0.97).

The HRs for fosphenytoin were larger for epilepsy, 1.62 (95% CI, 0.82–3.21. p = 0.14) and visual impairment, 1.87 (95% CI, 0.95–3.69 p = 0.07) than those of the other ASMs, but as the use of fosphenytoin was rare, the uncertainty related to the estimates was large and the results were not statistically significant.

### Multiple Anti-seizure Medications

The unadjusted Aalen-Johansen estimates indicated notable differences between outcome risks and the use of multiple ASMs compared to phenobarbital only, whereas the adjusted estimates did not (Figure 2). For example, after three years of follow-up, the adjusted cumulative incidences of cerebral palsy were 5.6% (95% CI, 3.2%-10.6%) for patients treated with phenobarbital and second-line ASMs, 4.6% (95% CI, 2.7%-7.3%) for patients treated with phenobarbital only and 0.66% (95% CI, 0.53%-0.81%) for patients with no ASM exposure. The corresponding unadjusted estimates were 35.5% (95% CI, 27.4%-43.6%), 17.1% (95% CI, 11.6%-22.7%) and 0.64% (95% CI, 0.51%-0.77%), respectively.

**Figure 2:**
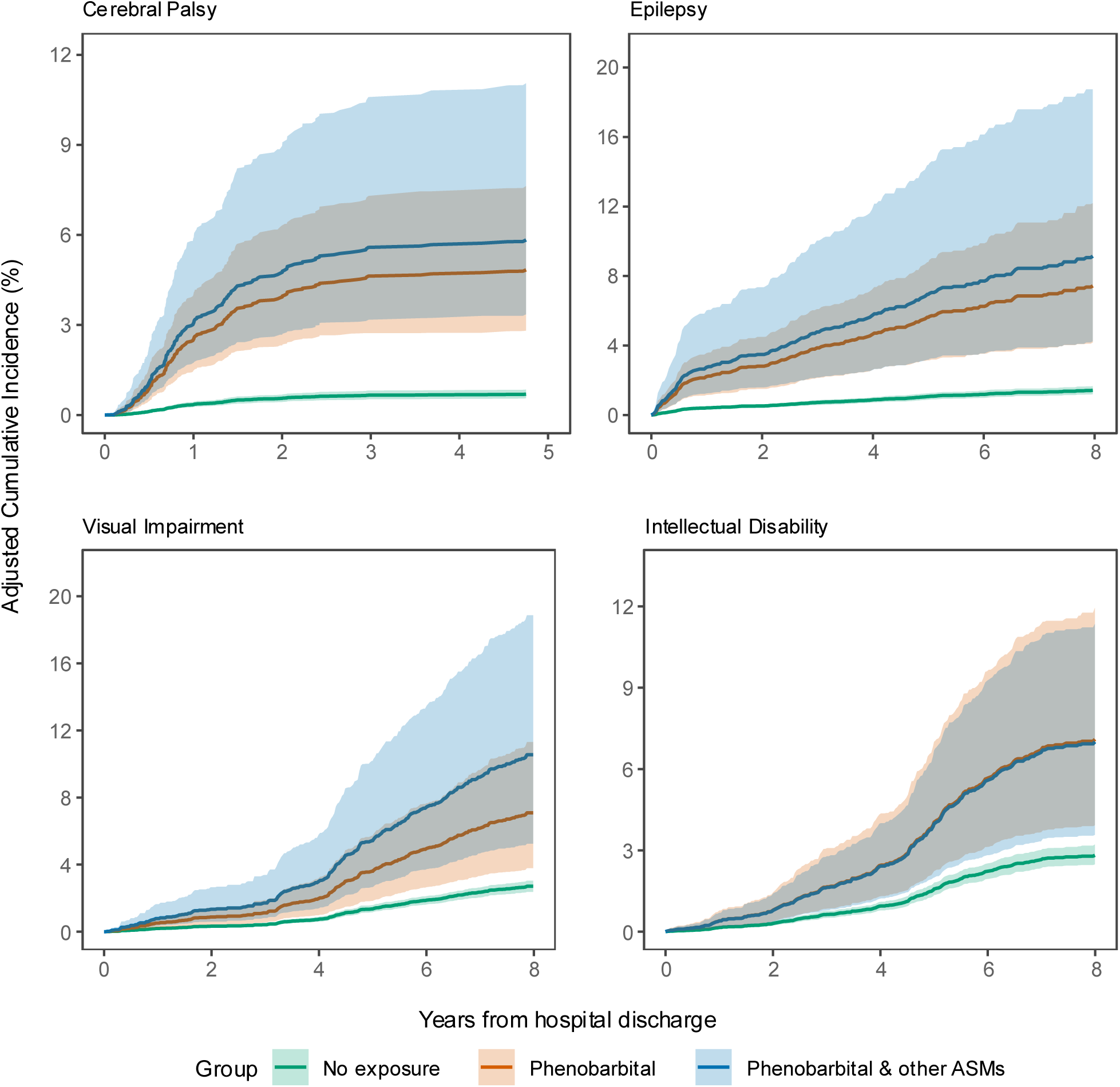
Adjusted cumulative incidences and 95 % bootstrap intervals based on 500 bootstrap samples for patients treated with phenobarbital and other ASMs. Alt Texts: Line plot showing adjusted cumulative incidences of cerebral palsy, epilepsy, visual impairment and intellectual disability as a function of time after hospital discharge in patients treated with phenobarbital or phenobarbital and other ASMs or no exposure to ASMs

Although patients exposed to both phenobarbital and other ASMs showed higher unadjusted incidences of cerebral palsy, epilepsy, visual impairment and intellectual disability, adjusted analyses did not provide evidence of worse outcomes after accounting for confounders and the uncertainty in the estimates.

### Phenobarbital Concentrations

Measured serum phenobarbital concentrations were available for 332 patients, enabling us to examine the relationship between concentration and clinical outcomes. In this subgroup, the hazard ratios for 50 µmol/l increase in maximum phenobarbital blood concentration associated with elevated hazard ratios for cerebral palsy (HR 1.48; 95% CI, 1.07–2.06, p = 0.02) and epilepsy (HR 1.64; 95% CI, 1.14–2.35, p = 0.007). No evidence for similar effect was found for visual impairment (HR 1.10; 95% CI, 0.71–1.70, p = 0.69) or intellectual disability (HR 1.08; 95% CI, 0.72–1.64, p = 0.71).

### Sensitivity Analysis

We evaluated the sensitivity of our results to: (1) excluding diagnoses classified as late by restricting the follow-up, (2) restricting the patient information to the neonatal period to eliminate large cumulative doses accumulated over longer hospitalization periods, and (3) assessed potential non-linear associations between the ASM use and the outcomes using restricted cubic splines. Restricting the follow-up did not affect the results but restricting the cumulative doses to the neonatal period or using restricted cubic splines increased the hazard ratios for phenobarbital (see Supplementary Material).

## Discussion

We found that exposure to phenobarbital in the neonatal intensive care unit was associated with elevated risk of adverse long term neurological outcomes, including epilepsy (HR 1.35; 95% CI, 1.11–1.62, p = 0.002), visual impairment (HR 1.20; 95% CI, 0.99–1.45, p = 0.06), and intellectual disability (HR 1.18; 95% CI, 0.99–1.41, p = 0.06). These findings are consistent with data from animal models and previous clinical studies^13,14^ and they reinforce concerns regarding potential long-term neurodevelopmental consequences of phenobarbital exposure in the neonatal period. Levetiracetam showed a more favorable profile, although it was still associated with a modest increase in risk, with hazard ratios around 1.1. In a recent systematic review, phenobarbital was concluded to be probably more effective than levetiracetam in achieving seizure control after first loading dose and after maximal loading dose, but data on mortality before discharge, mechanical ventilation and epilepsy post-discharge were uncertain.^3^ A recent meta-analysis on efficacy and safety of levetiracetam versus phenobarbital found no significant difference in the efficacies, but levetiracetam had a more favorable adverse effect profile than phenobarbital.^27^ Midazolam was also associated with lower hazard ratios for long term problems than phenobarbital, but its potent sedative effect often limits its use to third-line drug in treating neonatal seizures.

Although a significant relationship was observed between exposure to phenobarbital and neurologic outcomes, the sensitivity analyses showed that the cumulative dose did not offer extra insight into outcome risk for exposed patients. We did not consequently find evidence of a dose-dependent effect of phenobarbital (see Supplementary Material). This may reflect the relatively short courses administered, as the median cumulative dose was 28.4 mg/kg compared with a recommended initial loading dose of 20 mg/kg. This finding is consistent with the study by Saxena et al., in which infants receiving a loading dose of phenobarbital for seizure control and randomized to either short-term maintenance therapy or no maintenance therapy showed comparable risks of abnormal neurological assessment at 24 months of age.^28^

Increased maximum serum phenobarbital concentrations were associated with elevated hazard ratios for cerebral palsy and epilepsy. This could reflect a toxic effect after higher phenobarbital concentrations, while larger cumulative doses might not increase the risk as long as maximum concentrations remain at a lower level. Individual variation on phenobarbital pharmacokinetics could also play a role.^29^

This study has several limitations. The main problem pertains to distinguishing the effect of the underlying clinical problem that prompted seizures and ASM use from detrimental effect of the treatment. Previous studies have also struggled with the same problem. Typical etiologies (hypoxic-ischemic encephalopathy, ischemic stroke, and intracranial hemorrhage) are associated with both short and long term neurological problems and while we adjusted our analysis for these diagnoses, some effect may have persisted. The different models used in this study produced varying hazard ratios and we chose to present most conservative estimate. We believe that the comparisons between patients exposed to different ASMs may partly circumvent the problem. Still, phenobarbital use as a first-line treatment may have been skewed towards more benign clinical cases, and second- and third-line drugs to more severe cases, and we therefore addressed this by the multiple ASM analysis. Despite this potential bias, our study revealed that phenobarbital exhibited worse hazard ratios for cerebral palsy, epilepsy and visual impairment compared to levetiracetam or midazolam. Fosphenytoin was used too rarely for reliable analyses.

The reliability of our results is increased by the fact that the data are population-based and derived from a single center. The Helsinki University Hospital (HUS) catchment area encompasses approximately 1.7 million residents and 18,000 deliveries per year, with all infants requiring neonatal intensive care treated at the New Children’s Hospital NICU. Children requiring specialized pediatric or child neurological care are followed within HUS hospitals, and diagnostic data were available for 18,548 of 19,940 eligible children (93%).

## Conclusion

Using real-world data and long-term follow-up in a comprehensive population-based sample of infants treated with ASMs we showed exposure to phenobarbital to be associated with increased hazard ratios for epilepsy, visual impairment, and intellectual disability. These results align with previous findings linking phenobarbital to neurodevelopmental harm and emphasize the need for its cautious use in neonates. Levetiracetam had more favorable safety profile.

## Data Availability

All data produced in the present study are available upon reasonable request to the authors

## Conflict of Interest Disclosures (includes financial disclosures)

The authors have no conflicts of interest relevant to this article to disclose.

## Role of Funder/Sponsor (if any)

The Foundation for Pediatric Research and The Association of Friends of the University Children’s Hospitals had no role in the design and conduct of the study.

## Data Sharing Statement

Deidentified individual participant data will not be made available.

## Abbreviations

ASM: anti-seizure medication
HIE: hypoxic-ischemic encephalopathy
HR: hazard ratio
HUS: Helsinki University Hospital
IVH: intraventricular hemorrhage
NICU: neonatal intensive care unit

## What’s known on This Subject

Anti-seizure medications (ASM) are widely used in neonatal intensive care, but there is limited evidence for their long-term outcomes. Phenobarbital is the only ASM approved for use in neonates, but it has been linked with adverse effects. Other ASMs are used off-label in infants.

## What This Study Adds

By using real world data we showed increased hazard ratios for epilepsy, visual impairment and intellectual disability in infants exposed to phenobarbital. Although no dose-dependent relationship was observed, higher maximal plasma phenobarbital concentrations were associated with increased hazard ratios.

## Contributors Statement Page

Dr Markus Leskinen contributed to conceptualization of the study, performed literature searches and contributed to interpretation and writing (drafting and editing).

Volmar Kaipainen conducted data analyses, contributed to data curation, interpretation, software, visualization, and writing (drafting and editing).

Mine Ögretir contributed to data analyses, curation, software and writing (review and editing).

Drs Marjo Metsäranta, Matti Hero and Samuli Rautava contributed in conceptualization of the study and writing (review and editing).

Dr. Miika Koskinen M.K. conceptualized the study, supervised and administrated the project, interpreted and validated the data, acquired funding and critically reviewed and revised the manuscript.

All authors approved the final manuscript as submitted and agree to be accountable for all aspects of the work.

## Acknowledgments

This project received funding from institutional sources of HUS, The Association of Friends of the University Children’s Hospitals (Lastenklinikoiden Kummit ry) and The Foundation for Pediatric Research, Finland. We wish to acknowledge the support of Mona Halme, Mira Kilpi, and Mira Kuusinen from the Children and Adolescents’ Data Services Team at Helsinki University Hospital (HUS). ChatGPT Version 1.2025.350 and Microsoft Copilot were used in order to refine the language of the work.

## References

1. Gade C, Trolle S, Mørk M, et al. Massive presence of off_Jlabel medicines in Danish neonatal departments: A nationwide survey using national hospital purchase data. Pharmacol Res Perspect. 2023;11(1):e01037. doi:10.1002/prp2.1037

2. Barr J, Brenner-Zada G, Heiman E, et al. Unlicensed and Off-Label Medication Use in a Neonatal Intensive Care Unit: A Prospective Study. Am J Perinatol. 2002;19(2):067–072. doi:10.1055/s-2002-23557

3. Abiramalatha T, Thanigainathan S, Ramaswamy VV, Pressler R, Brigo F, Hartmann H. Anti_Jseizure medications for neonates with seizures. Cochrane Database Syst Rev. 2023;2023(10):CD014967. doi:10.1002/14651858.cd014967.pub2

4. Gettings JV, Soul JS. Updates in Neonatal Seizures. Clin Perinatol. 2025;52(2):375–393. doi:10.1016/j.clp.2025.02.008

5. Pressler RM, Abend NS, Auvin S, et al. Treatment of seizures in the neonate: Guidelines and consensus_Jbased recommendations—Special report from the ILAE Task Force on Neonatal Seizures. Epilepsia. 2023;64(10):2550–2570. doi:10.1111/epi.17745

6. Painter MJ, Scher MS, Stein AD, et al. Phenobarbital Compared with Phenytoin for the Treatment of Neonatal Seizures. N Engl J Med. 1999;341(7):485–489. doi:10.1056/nejm199908123410704

7. Sharpe C, Reiner GE, Davis SL, et al. Levetiracetam Versus Phenobarbital for Neonatal Seizures: A Randomized Controlled Trial. Pediatrics. 2020;145(6):e20193182. doi:10.1542/peds.2019-3182

8. Long D, Sutton C, Hale J. Efficacy of Levetiracetam vs Phenobarbital as First Line Therapy for the Treatment of Neonatal Seizures. J Pediatr Pharmacol Ther. 2024;29(5):482–486. doi:10.5863/1551-6776-29.5.482

9. Bättig L, Dünner C, Cserpan D, et al. Levetiracetam versus Phenobarbital for Neonatal Seizures: A Retrospective Cohort Study. Pediatr Neurol. 2023;138:62–70. doi:10.1016/j.pediatrneurol.2022.10.004

10. Kaushal S, Tamer Z, Opoku F, Forcelli PA. Anticonvulsant drug–induced cell death in the developing white matter of the rodent brain. Epilepsia. 2016;57(5):727–734. doi:10.1111/epi.13365

11. Bittigau P, Sifringer M, Genz K, et al. Antiepileptic drugs and apoptotic neurodegeneration in the developing brain. Proc Natl Acad Sci. 2002;99(23):15089–15094. doi:10.1073/pnas.222550499

12. Forcelli PA, Kim J, Kondratyev A, Gale K. Pattern of antiepileptic drug–induced cell death in limbic regions of the neonatal rat brain. Epilepsia. 2011;52(12):e207–e211. doi:10.1111/j.1528-1167.2011.03297.x

13. Falsaperla R, Mauceri L, Pavone P, et al. Short-Term Neurodevelopmental Outcome in Term Neonates Treated with Phenobarbital versus Levetiracetam: A Single-Center Experience. Behav Neurol. 2019;2019(1):1–8. doi:10.1155/2019/3683548

14. El-Dib M, Soul JS. The use of phenobarbital and other anti-seizure drugs in newborns.cSemin Fetal Neonatal Med. 2017;22(5):321–327. doi:10.1016/j.siny.2017.07.008

15. Farwell JR, Lee YJ, Hirtz DG, Sulzbacher SI, Ellenberg JH, Nelson KB. Phenobarbital for Febrile Seizures — Effects on Intelligence and on Seizure Recurrence. N Engl J Med. 1990;322(6):364–369. doi:10.1056/nejm199002083220604

16. Meyn DF, Ness J, Ambalavanan N, Carlo WA. Prophylactic Phenobarbital and Whole-Body Cooling for Neonatal Hypoxic-Ischemic Encephalopathy. J Pediatr. 2010;157(2):334–336. doi:10.1016/j.jpeds.2010.04.005

17. Donovan MD, Griffin BT, Kharoshankaya L, Cryan JF, Boylan GB. Pharmacotherapy for Neonatal Seizures: Current Knowledge and Future Perspectives. Drugs. 2016;76(6):647–661. doi:10.1007/s40265-016-0554-7

18. Maitre NL, Smolinsky C, Slaughter JC, Stark AR. Adverse neurodevelopmental outcomes after exposure to phenobarbital and levetiracetam for the treatment of neonatal seizures. J Perinatol. 2013;33(11):841–846. doi:10.1038/jp.2013.116

19. Hughes S, Blumenthal M, Johnson A, Limjoco J, Lim SY. Pharmacokinetics and Pharmacodynamics of Levetiracetam in Neonatal Seizures: What We Still Need to Know. J Pediatr Pharmacol Ther. 2025;30(2):170–181. doi:10.5863/1551-6776-30.2.170

20. Ahmad KA, Desai SJ, Bennett MM, et al. Changing antiepileptic drug use for seizures in US neonatal intensive care units from 2005 to 2014. J Perinatol. 2017;37(3):296–300. doi:10.1038/jp.2016.206

21. Zeller B, Giebe J. Pharmacologic Management of Neonatal Seizures. Neonatal Netw. 2015;34(4):239–244. doi:10.1891/0730-0832.34.4.239

22. Glass HC, Kan J, Bonifacio SL, Ferriero DM. Neonatal Seizures: Treatment Practices Among Term and Preterm Infants. Pediatr Neurol. 2012;46(2):111–115. doi:10.1016/j.pediatrneurol.2011.11.006

23. Ng E, Taddio A, Ohlsson^a^ A. Intravenous midazolam infusion for sedation of infants in the neonatal intensive care unit. Cochrane Database Syst Rev. 2017;1(1):CD002052. doi:10.1002/14651858.cd002052.pub3

24. Arya V, Ramji S. Midazolam sedation in mechanically ventilated newborns: a double blind randomized placebo controlled trial. Indian Pediatr. 2001;38(9):967–972.

25. Denz R, Klaaßen_JMielke R, Timmesfeld N. A comparison of different methods to adjust survival curves for confounders. Stat Med. 2023;42(10):1461–1479. doi:10.1002/sim.9681

26. Grambsch PM, Therneau TM. Proportional hazards tests and diagnostics based on weighted residuals. Biometrika. 1994;81(3):515–526. doi:10.1093/biomet/81.3.515

27. Salim N, Qadri SI, Chughtai MAI, et al. Efficacy and safety of levetiracetam versus phenobarbitone for neonatal seizures: A systemic review and meta-analysis. Clin Neurol Neurosurg. 2025;258:109155. doi:10.1016/j.clineuro.2025.109155

28. Saxena P, Singh A, Upadhyay A, Gupta P, Sharma S, Vishnubatla S. Effect of Withholding Phenobarbitone Maintenance in Neonatal Seizures: A Randomized Controlled Trial. Indian Pediatr. 2016;53:1069–1073.

29. Mamiya K, Hadama A, Yukawa E, et al. CYP2C19 polymorphism effect on phenobarbitone. Eur J Clin Pharmacol. 2000;55(11-12):821–825. doi:10.1007/s002280050703

