## Supplement for "Neonatal Anti-seizure Medication Exposure and Long-term Neurological Outcomes"

### Sensitivity Analyses

#### Unadjusted Estimates

**Table 2.1: Unadjusted hazard ratios and 95 % confidence intervals. Hazard ratios are reported for patients receiving the median positive cumulative dose for each medication, when the reference is a patient with no exposure.**

|  | **Cerebral Palsy** | **Epilepsy** | **Visual Impairment** | **Intellectual Disability** |
| --- | --- | --- | --- | --- |
| **Phenobarbital** | 1.94  [1.63, 2.31] | 1.80  [1.52, 2.14] | 1.51  [1.29 1.77] | 1.65  [1.39, 1.96] |
| **Levetiracetam** | 1.10  [1.01, 1.20] | 1.19  [1.11, 1.27] | 1.19  [1.12, 1.25] | 1.12  [1.02, 1.22] |
| **Midazolam** | 1.11  [1.04 1.18] | 1.12  [1.06, 1.19] | 1.07  [0.96, 1.20] | 1.19  [1.12, 1.26] |
| **Fosphenytoin** | 4.81  [2.41, 9.62 | 3.66  [1.8, 7.46] | 2.80  [1.56, 5.03] | 1.69  [0.77, 3.69] |
| Single model containing all four medications was used for each outcome. | | | | |

#### Restricted Follow-up

Administrative censoring times of 1830 (5.01 years), 3647 (9.97 years), 4128 (11.30 years) and 3148 days (8.62 years) were used for cerebral palsy, epilepsy, visual impairment and intellectual disability respectively, which correspond to the 95th percentile of event times observed in the data. The results are reported in Table 2.2 and are similar to those reported in the main analysis. For cerebral palsy, restricting this further to 1096 days (3 years) made little difference and is not reported here.

**Table 2.2: Hazard ratios and 95 % confidence intervals, when the follow up is restricted. Hazard ratios are reported for patients receiving the median positive cumulative dose for each medication, when the reference is a patient with no exposure.**

|  | **Cerebral Palsy¹** | **Epilepsy¹** | **Visual Impairment²** | **Intellectual Disability³** |
| --- | --- | --- | --- | --- |
| **Phenobarbital** | 1.131  [1.824, 1.552] | 1.324  [1.096, 1.600] | 1.212  [1.000, 1.467] | 1.145  [0.956, 1.374] |
| **Levetiracetam** | 1.119  [1.025, 1.222] | 1.142  [1.052, 1.239] | 1.180  [1.108, 1.256] | 1.079  [0.977, 1.192] |
| **Midazolam** | 1.002  [0.807, 1.243] | 1.062  [0.972, 1.161] | 0.987  [0.818, 1.191] | 1.075  [1.004, 1.151] |
| **Fosphenytoin** | 0.940  [0.367, 2.409] | 1.610  [0.811, 3.199] | 1.886  [0.960, 3.705] | 1.156  [0.509, 2.625] |
| 1 Adjusted for gestational age, birth weight, hospitalisation time, mode of delivery, gender, IVH, HIE, stroke and birth asphyxia  2 Adjusted for gestational age, birth weight, hospitalisation time, mode of delivery, gender, IVH, HIE, stroke, birth asphyxia and retinopathy of prematurity.  3 Adjusted for gestational age, birth weight, hospitalisation time, mode of delivery, gender, IVH, HIE and birth asphyxia. | | | | |

#### Cumulative Doses Within Neonatal Period

Calculating the cumulative doses for the neonatal period only had a notable impact on the effect of phenobarbital (Table 2.3). We hypothesized that this may reflect information loss regarding the duration of the hospitalisation period, as phenobarbital appeared to be prone to bias from improper covariate adjustments compared to other ASMs, which can be seen from the unadjusted estimates in Table 2.1.

**Table 2.3: Hazard ratios and 95 % confidence intervals, when the cumulative dose is calculated up to 28-days only. Hazard ratios are reported for patients receiving the median positive cumulative dose for each medication, when the reference is a patient with no exposure.**

|  | **Cerebral Palsy¹** | **Epilepsy¹** | **Visual Impairment²** | **Intellectual Disability³** |
| --- | --- | --- | --- | --- |
| **Phenobarbital** | 1.62  [1.18, 2.24] | 1.63  [1.22, 2.17] | 1.50  [1.20, 1.87] | 1.40  [1.02, 1.92] |
| **Levetiracetam** | 1.19  [1.07, 1.34] | 1.11  [0.98, 1.25] | 1.17  [1.03, 1.34] | 1.01  [0.87, 1.16] |
| **Midazolam** | 0.98  [0.69, 1.38] | 1.12  [1.01, 1.25] | 0.97  [0.77, 1.20] | 1.16  [1.07, 1.26] |
| **Fosphenytoin** | 0.81  [0.29, 2.26] | 1.60  [0.77, 3.33] | 1.97  [0.99, 3.90] | 0.98  [0.47, 2.10] |
| 1 Adjusted for gestational age, birth weight, hospitalisation time, mode of delivery, gender, IVH, HIE, stroke and birth asphyxia  2 Adjusted for gestational age, birth weight, hospitalisation time, mode of delivery, gender, IVH, HIE, stroke, birth asphyxia and retinopathy of prematurity.  3 Adjusted for gestational age, birth weight, hospitalisation time, mode of delivery, gender, IVH, HIE and birth asphyxia. | | | | |

#### Assessing Non-linear Effects

Due to the very small number of patients (37 patients) receiving fosphenytoin, its effect was assumed linear, whereas possible non-linearity for the other medications was assessed using restricted cubic splines with three knots placed at 0.1, 0.5 and 0.9 quantiles of positive dose values. Evidence for non-linearity was evaluated using three criteria: (1) statistical significance for the non-linear term in the Wald test, (2) improvement in Akaike information criterion over the model containing a linear term, and (3) the overall change of shape in the fitted relationship. In the absence of evidence for non-linearity, only a linear term was included to maintain model parsimony.

##### Phenobarbital

Phenobarbital was the only medication showing evidence for non-linearity, and modelling with restricted cubic splines showed a notable increase in risk compared to a linear term only. To assess this further, the models were re-specified to incorporate phenobarbital exposure as both a binary indicator (exposed vs unexposed) and a linear term for the cumulative dose. ^1–3^ The positive doses were centered around the median dose, so that the exponentiated binary term represents the hazard ratio for exposure to the median dose, and the continuous term the hazard ratio associated with deviations from the median. This approach was adopted to address potential artifacts arising from zero-inflation of the exposure, as it calculates the dose-response relationship for the exposed patients only.

Under this specification, the overall effect of phenobarbital remained similarly elevated, but the dose component did not suggest any meaningful dose-dependent effects: HR 0.99 (95% CI, 0.98–1.00), HR 1.00 (95% CI, 0.99–1.01), HR 1.00 (95% CI, 0.99–1.01), and HR 1.00 (95% CI, 0.99–1.01) for cerebral palsy, epilepsy, visual impairment and intellectual disability respectively. Among exposed patients, the level of the dose did not provide any additional information about outcome risk, and the estimated associations were carried entirely by the binary exposure indicator (Table 2.4).

The magnitude of this effect was large and raises the possibility of confounding by indication, particularly as phenobarbital appeared more sensitive to imperfect covariate adjustment than other medications in earlier sensitivity analyses. It is therefore plausible that the binary exposure term reflects residual differences in underlying health status—such as HIE, IVH and seizures—or that the models were unable to allocate risk accurately between these conditions and treatment, as both were relatively rare.

We ultimately modelled phenobarbital using a single linear term, as it appeared less susceptible to the previously discussed issues and yielded estimates comparable to those for other ASMs and previous studies.

**Table 2.4: Hazard ratios and 95 % confidence intervals under different parametrization for phenobarbital. Hazard ratios are reported for patients receiving the median positive cumulative dose for each medication, when the reference is a patient with no exposure.**

|  | **Cerebral Palsy¹** | **Epilepsy¹** | **Visual Impairment²** | **Intellectual Disability³** |
| --- | --- | --- | --- | --- |
| **Phenobarbital** | 6.76  [3.84, 11.89] | 4.57  [2.57, 8.23] | 2.78  [1.60, 4.81] | 2.18  [1.23, 3.85] |
| **Levetiracetam** | 1.11  [1.03, 1.19] | 1.19  [1.03, 1.21] | 1.21  [1.13, 1.29] | 1.07  [0.95, 1.21] |
| **Midazolam** | 1.06  [0.99, 1.13] | 1.06  [0.99, 1.14] | 1.02  [0.88, 1.19] | 1.14  [1.07, 1.21] |
| **Fosphenytoin** | 1.16  [0.62, 2.01] | 1.58  [0.90, 2.78] | 1.84  [1.01, 3.33] | 1.10  [0.53, 2.27] |
| 1 Adjusted for gestational age, birth weight, hospitalisation time, mode of delivery, gender, IVH, HIE, stroke and birth asphyxia  2 Adjusted for gestational age, birth weight, hospitalisation time, mode of delivery, gender, IVH, HIE, stroke, birth asphyxia and retinopathy of prematurity.  3 Adjusted for gestational age, birth weight, hospitalisation time, mode of delivery, gender, IVH, HIE and birth asphyxia. | | | | |

1 Hosmer DW, Lemeshow S, Sturdivant RX. Applied Logistic Regression. *Wiley Ser Probab Stat* 2013; : 377–457.

2 Lorenz E, Jenkner C, Sauerbrei W, Becher H. Modeling Variables With a Spike at Zero: Examples and Practical Recommendations. *Am J Epidemiology* 2017; **185**: 650–60.

3 Sauerbrei W, Perperoglou A, Schmid M, *et al.* State of the art in selection of variables and functional forms in multivariable analysis—outstanding issues. *Diagn Progn Res* 2020; **4**: 3.
